# Changes in prediction modelling in biomedicine – do systematic reviews indicate whether there is any trend towards larger data sets and machine learning methods?

**DOI:** 10.1101/2024.08.09.24311759

**Authors:** Lara Lusa, Franziska Kappenberg, Gary S. Collins, Matthias Schmid, Willi Sauerbrei, Jörg Rahnenführer, the Topic Group for High-dimensional data of the STRATOS initiative

## Abstract

The number of prediction models proposed in the biomedical literature has been growing year on year. In the last few years there has been an increasing attention to the changes occurring in the prediction modeling landscape. It is suggested that machine learning techniques are becoming more popular to develop prediction models to exploit complex data structures, higher-dimensional predictor spaces, very large number of participants, heterogeneous subgroups, with the ability to capture higher-order interactions.

We examine these changes in modelling practices by investigating a selection of systematic reviews on prediction models published in the biomedical literature. We selected systematic reviews published since 2020 which included at least 50 prediction models. Information was extracted guided by the CHARMS checklist. Time trends were explored using the models published since 2005.

We identified 8 reviews, which included 1448 prediction models published in 887 papers. The average number of study participants and outcome events increased considerably between 2015 and 2019, but remained stable afterwards. The number of candidate and final predictors did not noticeably increase over the study period, with a few recent studies using very large numbers of predictors. Internal validation and reporting of discrimination measures became more common, but assessing calibration and carrying out external validation were less common. Information about missing values was not reported in about half of the papers, however the use of imputation methods increased. There was no sign of an increase in using of machine learning methods. Overall, most of the findings were heterogeneous across reviews.

Our findings indicate that changes in the prediction modeling landscape in biomedicine are less dramatic than expected and that poor reporting is still common; adherence to well established best practice recommendations from the traditional biostatistics literature is still needed. For machine learning best practice recommendations are still missing, whereas such recommendations are available in the traditional biostatistics literature, but adherence is still inadequate.

## 1 Introduction

Models that provide predictions are an important tool in diagnosis, prognosis and treatment selection for human diseases. Clinical prediction models estimate an individual’s risk of a specific health outcome, using known characteristics, typically demographic and medical information. The interest in prediction models in medicine is growing: in 2023, for example, about one of 25 papers indexed in PubMed could be retrieved searching for “predictive model” or “prediction model”, a number that is more than 2 times larger compared to twenty years earlier (https://esperr.github.io/pubmed-by-year/).

Despite the increase in prediction model studies, few of the developed models are implemented in clinical practice [1, 2]. Contibuting to the poor uptake is likely the poor adherence to methodological recommendations in the development of the models [1, 3, 4], which was also the main finding of the review of prediction models published in high-impact journals in 2008 [5]. Editorials and review papers relate the poor applicability to the increase in the number of publications that use large datasets (often derived from routinely collected data) and the widespread use of machine learning (ML) methods [6–15]. ML methods can be particularly complex and thus more prone to overfitting and are rarely validated using independent data [7]; often described as lacking transparency compared to predictions based on regression approaches [7], using limited subject matter expertise and providing models where the contribution of different predictors is difficult to interpret [6]. Particular types of large datasets are often described as commonly lacking sufficient quality and detail to answer clinically relevant questions or guide decision making [16]; the need to address many methodological issues before potentially useful prediction models can be developed using big data or routinely collected data has been stressed; these methodological issues include heterogeneity between populations, changes over time, differences across centers, under-representation of populations, missing data, lack of structure, inaccuracies, lack of calibration and insufficient data sharing [10, 11, 14].

Because of the changes in the type and availability of data and type of analysis strategies being used, many suggestions from the literature indicate that the existing best practice recommendations for design, conduct, analysis, reporting, impact assessment, and clinical implementation from the biostatistics and medical statistics literature are no longer sufficient alone to guide the use of prediction models when machine learning methods are being used [6, 10, 17–21]. Consequently, many initiatives have been launched to propose new guidelines for the development, reporting and critical appraisal of prediction models based on machine learning/artificial intelligence (ML/AI) methods [10]; these include the TRIPOD (for model development/validation), CONSORT (trials of AI interventions), SPIRIT (protocols of trials of AI interventions), and PROBAST (risk of bias assessment) guidelines and tool for ML/AI that were updated or are in development [19, 22–25].

The aim of this paper is to explore if and how the prediction modelling landscape is changing. We focus on prognostic models that have been developed for the prediction of a future health outcome event based on a model that uses multiple predictors [26].

Systematic reviews are a valuable tool for obtaining information about existing prognostic models, summarizing their predictive performance and quality, and information about the predictors used [27]. The number of systematic reviews on prognostic models in the biomedical literature in the last years raised at a pace comparable to the increase observed in the number of publications that develop or apply prognostic models. Systematic reviews are often focused on specific outcomes and target populations, including relatively few prediction models. In exceptional cases, such as the recent Covid review [4], the findings from hundreds of prediction models are described.

We explored whether the landscape of prediction model studies is changing by reviewing systematic reviews of prognostic models. We focused on 8 reviews published (or updated) in 2020-22 and examined in detail the characteristics of the prognostic models included in the eligible [27]; the characteristics include (but are not limited to) the number of study participants, number of candidate and final predictors, type and number of prediction models and measures that quantify the performance of the models. We focused only on model development and omitted models where only the results from an external validation of an existing model, without model development, are reported.

In the Methods section we explain how the reviews were selected and describe their characteristics in detail. In the Results section we summarize the findings, focusing on exploring any time trends, and conclude with the Discussion section.

## 2 Methods

### 2.1 Selection of the reviews

The initial search of systematic reviews was based on a manually curated list made publicly available by Gary Collins (https://twitter.com/GSCollins/status/1506249323180437507). The list included about 260 systematic reviews of prediction models published in various medical fields; the number of reviewed models ranged from 3 to 1382, 52 systematic reviews reviewed more than 50 models, about a half included fewer than 20 models. The reviews were published between 2004 and 2022, most of them in the 2010-2020 period.

We examined the list of the 260 reviews and screened the content of the 19 systematic reviews that were reported to include at least 50 models and were published in 2020-22. We excluded the reviews

- that could not be retrieved as full text (1 systematic review)
- for which the individual per paper/per model data were not available (or at least not as a table, 6 systematic reviews)
- that described less than 30 papers where prognostic models were developed (6 systematic reviews, 1 included only validation of models).

which led to 6 eligible systematic reviews.

Manual screening of the results of a PubMed search for systematic reviews of prognostic models identified two additional systematic reviews that were eligible for inclusion. External validation only studies were excluded from our analyses. Thus, in total 8 reviews paper (6 from the manually curated list, 2 from additional searching) were included in our review of the reviews. The selection process is displayed in Figure 1.

**Fig 1.**
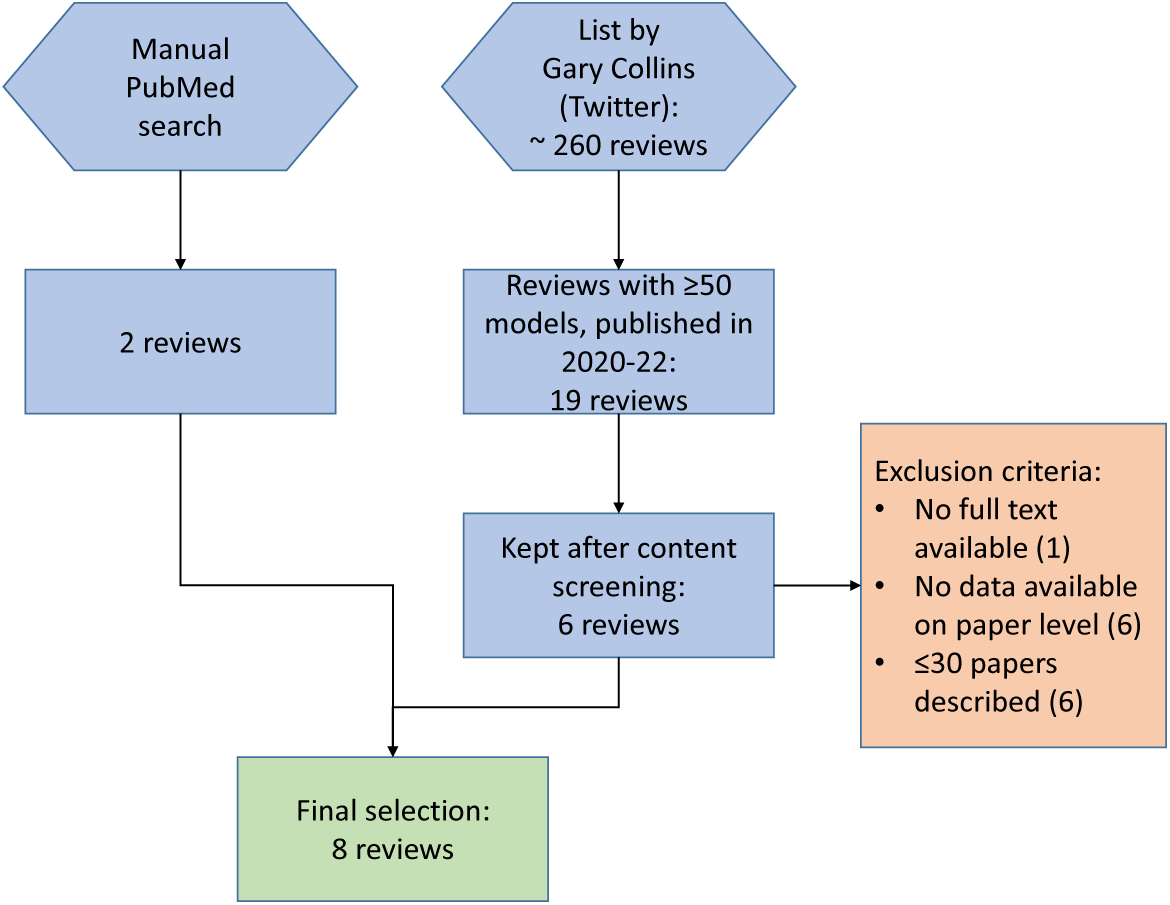
Flowchart displaying the process for selecting the reviews.

The main characteristics of the reviews were summarized using the PICOTS system [28] (**P**opulation, **I**ndex model, **C**omparator model, **O**utcome(s), **T**iming, **S**ettings), where we omitted the comparator model and reported only the timing related to the moment in time when the models are to be used in clinical practice.

We summarized the number of papers/models that were analyzed in each review, the time range of publication of the papers that they included and what type of information was available for each review.

For each paper/model we extracted information following the CHARMS checklist [29], including the number of study participants, the type of outcomes being predicted, the (candidate) predictors, analytical details (e.g., type of model, handling of missing data, selection of predictors), and evaluation of the model performance (discrimination, calibration, classification). The complete list of extracted data is available in the Supplementary table 1.

### 2.2 Data management

We organized the raw extracted data from each review in one table. Data were then processed manually and harmonized, where applicable, summarizing them to the categories considered later in the analysis. Only the information provided in the reviews was considered and we did not check and re-extract the original papers.

Some papers included in the reviews described more than one model. Our analyses were performed on a ‘per-paper’ basis, if not otherwise noted; this was done to avoid giving excessive weight to the papers that developed many different prediction models. Numerical data from different models described in the same paper were summarized using mean values (omitting missing values). A method or measure was considered as having been used/reported in the paper if it was used/reported for at least one of the models described in the paper.

For the type of prediction models, in a first step, we assigned the described methods to one of the following classes: ‘Neural network’, ‘Random Forest’, ‘Other tree-based’, ‘SVM’ (Support vector machine), ‘Boosting’, and ‘Other (ML)’, ‘(Penalized) Logistic Regression’, ‘Linear Regression’, ‘Cox Regression’, ‘Other (Stats)’; the categories‘NA/Not reported/Unclear’ and ‘Other’ (not clear if statistical or ML) were also used. The class ‘Tree-based’ refers to single trees only, not to random forests, tree-based boosting approaches or any other ensemble methods.

In the review from Li [30], where the information was given only on a per-paper basis, a list of used prediction methods was provided. These were classified as ‘Multiple (ML)’ or ‘Multiple (Both)’, as appropriate; statistical models were never used exclusively in this review.

Also for the other reviews, we defined for each paper if the models were developed using exclusively statistical methods (‘(Penalized) Logistic Regression’, ‘Linear Regression’, ‘Cox Regression’, ‘Other (Stats)’), exclusively ML methods ( ‘Neural net’, ‘Random Forest’, ‘Tree-based’, ‘SVM’ (Support vector machine), ‘Boosting’, and ‘Other (ML)’), or both, or if the information was unclear (‘NA/Not reported/Unclear’ and ‘Other).

The measures that quantify the predictive performance for internal validation were grouped into the three categories ‘Discrimination’, ‘Calibration’, and ‘Classification’ (as suggested in the CHARMS checklist [29]). The area under the receiver-operator characteristic curve (AUC or AUROC, sometimes also just denoted as ROC) and the C-index (sometimes C-statistic) were considered to be measures for discrimination. Calibration plots (i.e. observed vs expected risks) and calibration slopes, calibration in-the-large, Hosmer-Lemeshow tests, Greenwood-D’Agostino-Nam tests, and Gronnesby and Borgan tests were all categorized as ‘Calibration’ measures. Finally, the group of ‘Classification’ measures entailed Accuracy, Sensitivity (or Recall), Specificity, Positive Predictive Value (or Precision), Negative Predictive Value, F_1_-score, Youden-index, Positive Likelihood Ratio, Negative Likelihood Ratio, and the Diagnostic Odds Ratio.

Internal validation methods were grouped in categories: cross-validation, bootstrap (including resampling or jacknife), split-sample (random, temporal or other), other (not specified or combinations), or missing information (NA); external validation methods were not further categorized, as the information was very limited.

The handling of missing values was evaluated at per-model level and categorized as: predictor omission, missing indicator methods/Dummy, Complete Case, Single imputation, Multiple imputation, Other imputation, Unclear/No information, Other, No Need To Report/None.

### 2.3 Presentation of the results

We summarized the characteristics of the papers/models that were reviewed, overall and stratified by systematic review, in order to account for heterogeneity of the reviews into account.

Several graphical displays were used. Overall time trends were displayed using scatterplots with an added smoothing line obtained with a loess smoother with 95% confidence bands (using the default settings of the geom smooth() function from the ggplot2 R package). Summaries of numerical variables were displayed with a combination of violin plots and boxplots, to display the summary statistics and the overall distribution of the data. Scatterplots were used to compare the number of candidate and of final predictors, where the sizes of the individual dots reflected the respective underlying frequency. Categorical variables were summarized by stacked barplots (with absolute and relative numbers). The occurrence of the different types of measures was displayed using Sankey plots.

In tables the numerical variables were summarized using median (med), arithmetic mean (mean), the interval between minimum and maximum (range), and interquartile range (IQR). Categorical variables were summarized using frequencies and percentages.

All analyses were conducted in the statistical programming software R, version 4.2.2 [31]. For the display of the results, the R packages ggplot2 [32], ggalluvial [33], ggpubr [34], and ggh4x [35] were used.

### 2.4 Initial data analysis

Decisions about how to present data were based on initial data analysis (IDA), where the distributions of the variables were explored using descriptive statistics [36].

IDA indicated the exclusion of papers published before 2005, due to their small number (n=27). To explore the time trends in some analyses we grouped the year of model publication into intervals with the following intervals 2005-2009, 2010-2014, 2015-2019 and 2020-2021. IDA also indicated the removal of the review of Wynants et al. on COVID-19 [4] from the time trend analysis, as this review contains 75% of the papers included in our review in the 2020/21 period and it would have dominated the results in the 2020/21 period. The results of the Wynants review [4] were therefore included in the overall analyses and commented on separately in the time trend analyses.

Some information about the type of the outcome (binary, categorical, time to event, numerical) is available indirectly, by examining the type of models used; however, in most reviews the information was not reported explicitly. For this reason we did not exclude numerical outcomes from the analysis of the number of outcome events. We decided not to analyze the time trends of the number of outcome events per predictor, as the information was very sparse and dominated by the information provided in Wynants [4].

We decided not to analyze data on clinical utility of the prognostic modes, as this information was rarely reported in the reviews.

## 3 Results

Here we describe the main characteristics of the reviews and the characteristics that were selected for our analyses, analyzing complete data, stratifying the results per review, and summarizing the time trends.

The TRIPOD (Transparent Reporting of a multivariable prediction model for Individual Prognosis Or Diagnosis) statement gives a set of recommendations for the reporting of studies involving the development or validation of a prediction model [22]. This statement is referenced in broad terms in three reviews (Wynants [4], Ogink [37], He [38]), and not considered at all in two reviews (Ndjaboue [39], Sun [40]). For the remaining three reviews (Li [30], Haller [41], Gade [42]), the information about adherence to the TRIPOD statement is given on an overall basis in Li [30] and

Haller [41], and for each paper individually in Gade [42].

### 3.1 Main characteristics of the reviews

We included 8 systematic reviews that described the prediction models in different medical fields (COVID-19 by Wynants et al. [4], vascular surgery by Li et al. [30], heart failure by Sun et al. [40], diabetes by Ndjaboune et al. [39], orthopaedics by Ogink et al. [37], cervical cancer by He et al. [38], organ transplantation by Haller et al. [41], falls by Gade et al. [42]); the PICOTS elements are described in Table 1. Five reviews (Wynants [4], Ndjaboue [39], Haller [41], He [38], Gade [42]) considered all available prognostic models up to the time of search, two studies focused on machine learning based prediction models (selecting studies that included at least one ML-based prediction, Ogink [37] and Li [30]), and one study considered models published in the previous ten years (Sun [40]). Only one review considered the study design as an inclusion criteria (Gade [42], including only prospective cohort studies); the study design was reported in two additional reviews: Haller [41] reported only cohort studies, observational studies were the majority for Wynants [4], which included also some registry studies. The raw information was provided at per-paper level only in one review (Li [30]), and per-model in the other reviews. In the Wynants review [4] 116/501 models used some type of imaging techniques (mostly CT scans or X-ray), and one review (Li [30]) included also prediction models for image segmentation (55/215 papers, which were included in the analyses as there was no indication that the aim of the analysis was not prognostic).

**Table 1.** PICOTS elements of the selected reviews. - Population, Index model, Comparator model (omitted), Outome(s), Timing, Settings.

| Review | Population | Index model | Outcome | Timing of use | Settings |
| --- | --- | --- | --- | --- | --- |
| Wynants et al. [4] | Patient with confirmed COVID-19 | All available prognostic models | All outcomes | Moment of COVID diagnosis | Inpatients and outpatients |
| Li et al. [30] | Patients with vascular conditions | Prediction models that use ML methods (prognosis, diagnosis and segmentation) | All outcomes | Not specified | Not specified |
| Sun et al. [40] | Patient with heart failure | All available prognostic models | All-cause mortality or all-cause readmission of heart failure patients | Not specified | Inpatients and outpatients |
| Ndjaboue et al. [39] | People of any age with pre-diabetes and any type of diabetes, except gestational diabetes | All available models with reported internal and/or external validation | Diabetes-related health conditions, mortality and mental health | Not specified | All settings |
| Ogink et al. [37] | Surgical orthopaedic population | Prognostic models from studies that included at least one ML-based prediction | Orthopaedic surgical outcomes | Intra-operative and post-operative | All settings |
| He et al. [38] | Patients diagnosed with cervical cancer | All available models (with at least two predictors) | Clinical outcome (recurrence, metastasis, death, etc.) | Not specified | All settings |
| Haller et al. [41] | Recipients or donors in living organ transplantation | All available models (with at least two predictors) | Any outcome occurring after transplantation/donation in the recipient or donor | Counseling | All settings |
| Gade et al. [42] | Community-dwelling older adults (60+) of the general population | All available models (with at least two predictors) | Falls (defined as unexpected event in which the patients come to rest on the ground, floor or lower level) | No restriction | General population |

Some information was systematically missing for some reviews, and there were missing values also when the information was intended to be summarized in the review, indicating that some of the reviewed papers did not provide all the information (Supplementary file 1). For example, the number of outcome events or the number of candidate predictors was often missing, making the analysis of the number of outcome events per variable problematic. Information for both number of outcome events and number of candidate predictors was available for only three reviews, which all also directly provided the number of event per variable, however with many missing values. The heterogeneity across reviews is further addressed in Subsection 3.2.

We identified and excluded 363 models that were included in the reviews only for validation purposes and 51 papers that were published before 2005 (published during 1987-2004). Overall, in our analyses we considered 887 papers and 1448 models from the 8 systematic reviews; the number of papers included in each review ranged from 28 to 368, the number of models from 49 to 501 (Table 2).

**Table 2.** Overall numbers of models and papers included and excluded from the analyses, by review.

| Review | Field | Time | Included |  | Excluded |  |
| --- | --- | --- | --- | --- | --- | --- |
|  |  |  | Models | Papers | Pure valida-<br>tion models | Papers pub-<br>lished before<br>2005 |
| Wynants | Covid-19 | 2020 to 2022 | 501 | 368 | 230 | 0 |
| Li | Vascular Surgery | 1991 to 2021 | - | 202 | 0 | 10 |
| Sun | Heart Failure | 2011 to 2021 | 176 | 78 | 104 | 0 |
| Ndjaboue | Diabetes | 2000 to 2020 | 175 | 75 | 0 | 5 |
| Ogink | Orthopaedic | 1996 to 2020 | 218 | 56 | 0 | 16 |
| He | Cervical cancer | 1987 to 2020 | 74 | 52 | 27 | 3 |
| Haller | Organ Transplantation | 2004 to 2021 | 48 | 35 | 0 | 1 |
| Gade | Falls | 1994 to 2019 | 54 | 21 | 2 | 16 |

### 3.2 Overall results and time trends

In this section we report the overall results (based on all included papers), describe time trends and summarize separately the papers included in the Wynants (COVID) paper. The descriptive statistics are also stratified by review. Unless otherwise noted, the summaries are given at per-paper level.

The number of papers included in the systematic reviews was larger in the more recent years (Figure 3 and Supplementary file 1 for additional information by review). The papers published in 2020/21 represented the majority of the papers (40% were from the Wynants review and 14% from the other reviews), while the 2005/09 period was the least represented with 46 papers (5%). The increase in the number of papers was consistent across reviews (Supplementary file 1).

#### 3.2.1 Number of study participants

The number of study participants was reported in 7 reviews (88% of papers). The overall distribution of the number of study participants (all reviews considered jointly) was strongly right-skewed (med=395, mean=17511); the number was above 200,000 in 10 papers/models, all of which were published in 2019 or later. The number of study participants included in the papers/models varied: the median values ranged from 200 (Li [30]) to 5460 (Ogink [37]) (Supplementary file 1 for additional overall and per review summaries).

Overall, the number of study participants increased over time as did the percentage of papers/models for which the information was available from the reviews (complete data were used in Figure 2, years were grouped in Figure 3 in Supplementary file 1). In the Wynants [4] review the number of study participants was considerably smaller compared to the papers included in the other reviews and published over the same period (2020/21) (med: 365 vs 660, Supplementary file 1).

**Fig 2.**
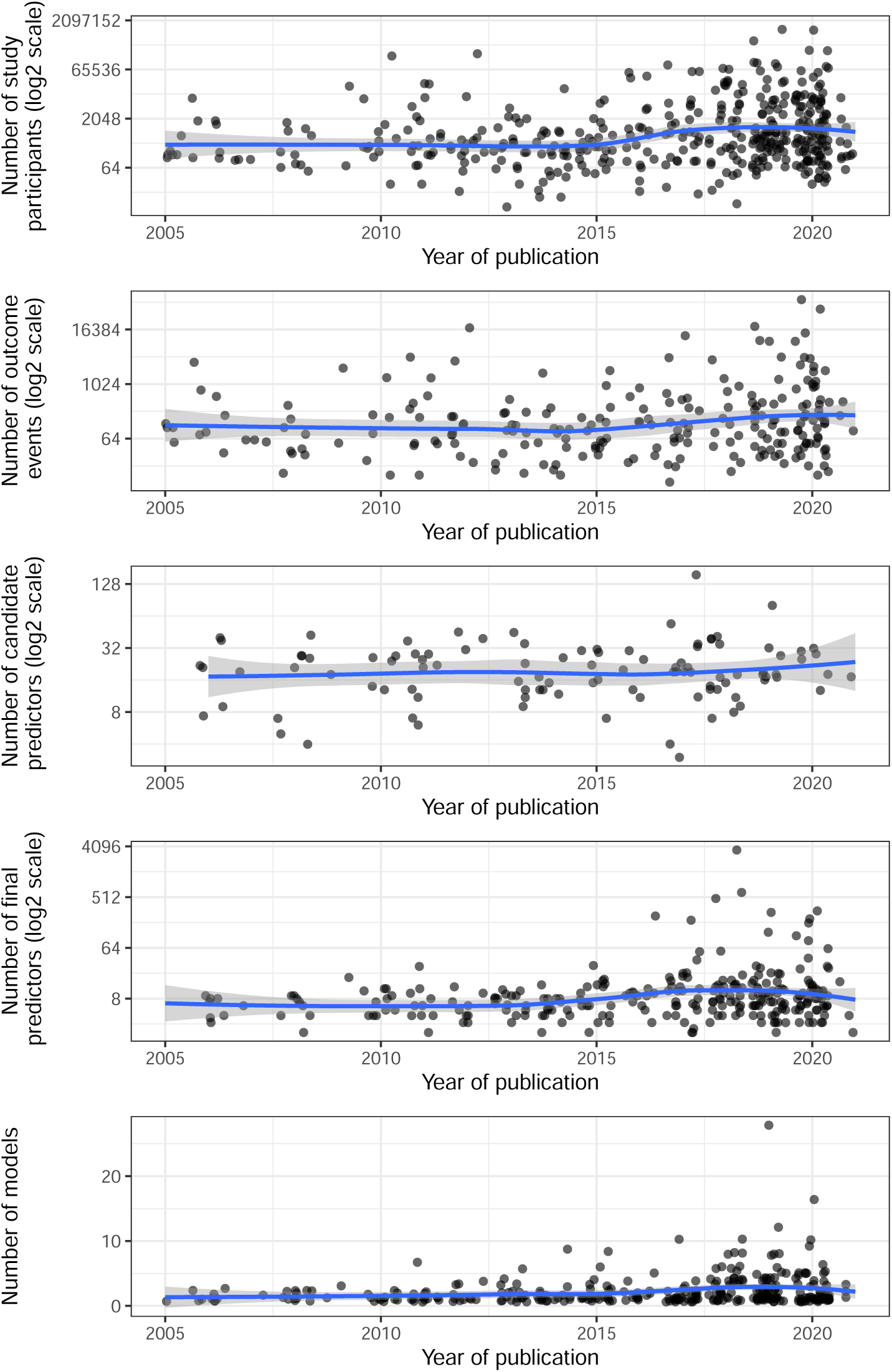
Time trends for number of study participants, events, candidate predictors, final predictors and models. The data from Wynants were not included. Each dot represents one paper; the blue trend lines represent the overall associations and are obtained using a loess smoother; the gray ribbons are 95% confidence bands.

**Fig 3.**
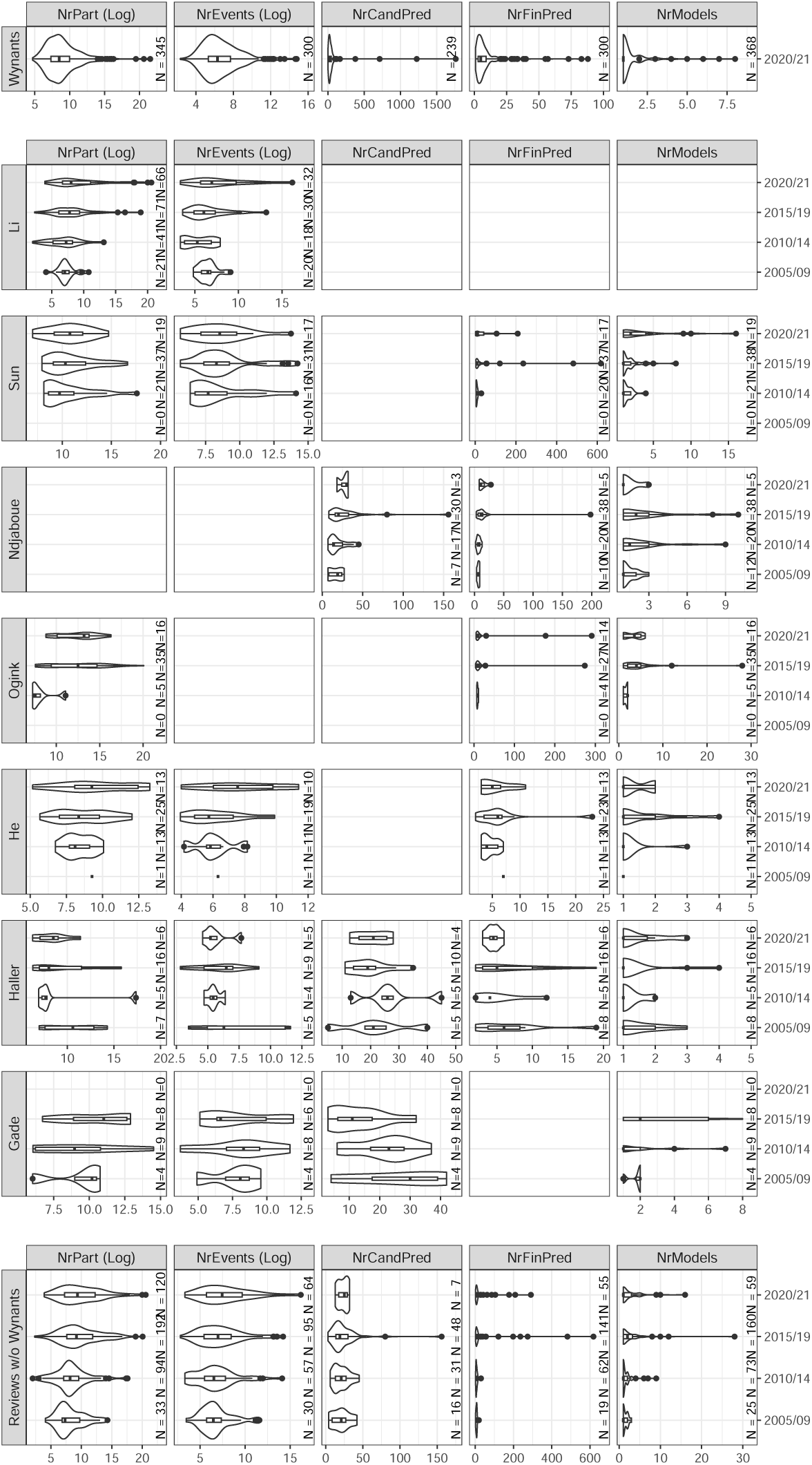
Descriptive characteristics by review and grouped by the intervals for year of publication. The displayed variables are: number of models (NrModels), number of study participants (NrPart, log2 scale), number of outcome events (NrEvents, log2 scale), number of candidate predictors (NrCandPred), and number of final predictors (NrFinPred). Results are shown with individual x-axes. Some values were omitted from this plot to better visualize the relevant areas: for NrCandPred, a value of 3463 for Ndjaboue, year 2015/19, and a value of 15000 for Wynants were not shown. For NrUsedPred, a value of 3512 for Sun, year 2015/19, a value of 1302 for Sun, year 2020/21, and a value of 483.3 for Wynants were not shown.

#### 3.2.2 Number of outcome events

The number of outcome events was reported in 6 reviews (62% of all papers); missing number of outcome events were for the reviews that, in principle, reported the information (Figure 3 and Supplementary file 1) and they were present for all types of outcomes. For example, the number of outcome events was missing for 47/326 models that used (penalized) logistic regression and for 50/180 for models that used Cox regression. The distribution of the number of outcome events was strongly right-skewed (med=89, mean=822, range: 5 to 74661), with considerable variability across reviews (from med=53 in Haller [41] to med=298 in Sun [40], Figure 3 and Supplementary file 1).

The number of outcome events increased over time (summaries in Figure 2 and Figure 3 and in Supplementary file 1). Within the reviews of Li [30] and Sun [40], where the largest number of models developed in different years were included, the increase during the 2010s was noticeable (Figure 3). Similarly as for the number of study participants, very large numbers of events were used mostly in models that were published more recently (out of the 9 papers that reported more than 10,000 events, one was published each in 2012, 2017, and 2019, and 6 in 2020). The Wynants review [4] described papers with fewer events compared to the other papers from the same period, and reported the information more frequently than the other reviews.

#### 3.2.3 Number of predictor variables (candidate and final)

The number of candidate predictors was available in 4 reviews (38% of papers) and the number of final predictors (i.e., those in the final model) in 5 reviews (65% of papers); one review provided only the number of candidate predictors (Gade [42]) and two only the number of final predictors. Among the reviews that collected information on the number of predictors, the information about candidate predictors was often missing, while the number of final predictors was reported most of the times (Supplementary file 1).

The overall median number of candidate predictors was 25 (mean=84, IQR = 14 to 40), the median number of final predictors was 6 (mean=21, IQR = 4 to 11); the distribution of the number of candidate predictors was strongly right-skewed, with mean values much larger than medians in the most recent papers. Over time the number of candidate predictors remained rather stable; it increased only in one review (Ndjaboue [39]).

The mean number of final predictors increased over time but the median values remained rather stable, two reviews showed a marked increase (Ndjaboue [39] and Sun [40]); most studies with very large number of predictors appeared after 2015 (Figure 3 and Supplementary file 1). The papers included in the Wynants review [4] used fewer predictors compared to the papers from the other reviews analysed in this work and published in the same period (2020/21) (med: 4 vs 8, mean: 11 vs 25), in the other reviews the information on candidate predictors in the 2020/21 papers was too scarce (n=7) to make meaningful comparisons. For the Wynants review [4] we compared the number of candidate and final predictors in models that used imaging with the other models, and observed that the number of candidate predictors was larger in models with imaging data (med: 112 vs 34, mean: 80 vs 46) while the number of final predictors was smaller (med: 4 vs 7, mean: 15 vs 23).

A subset of studies used all available predictors, but generally the number of final predictors was greatly reduced by some type of predictor selection (Figure 4).

**Fig 4.**
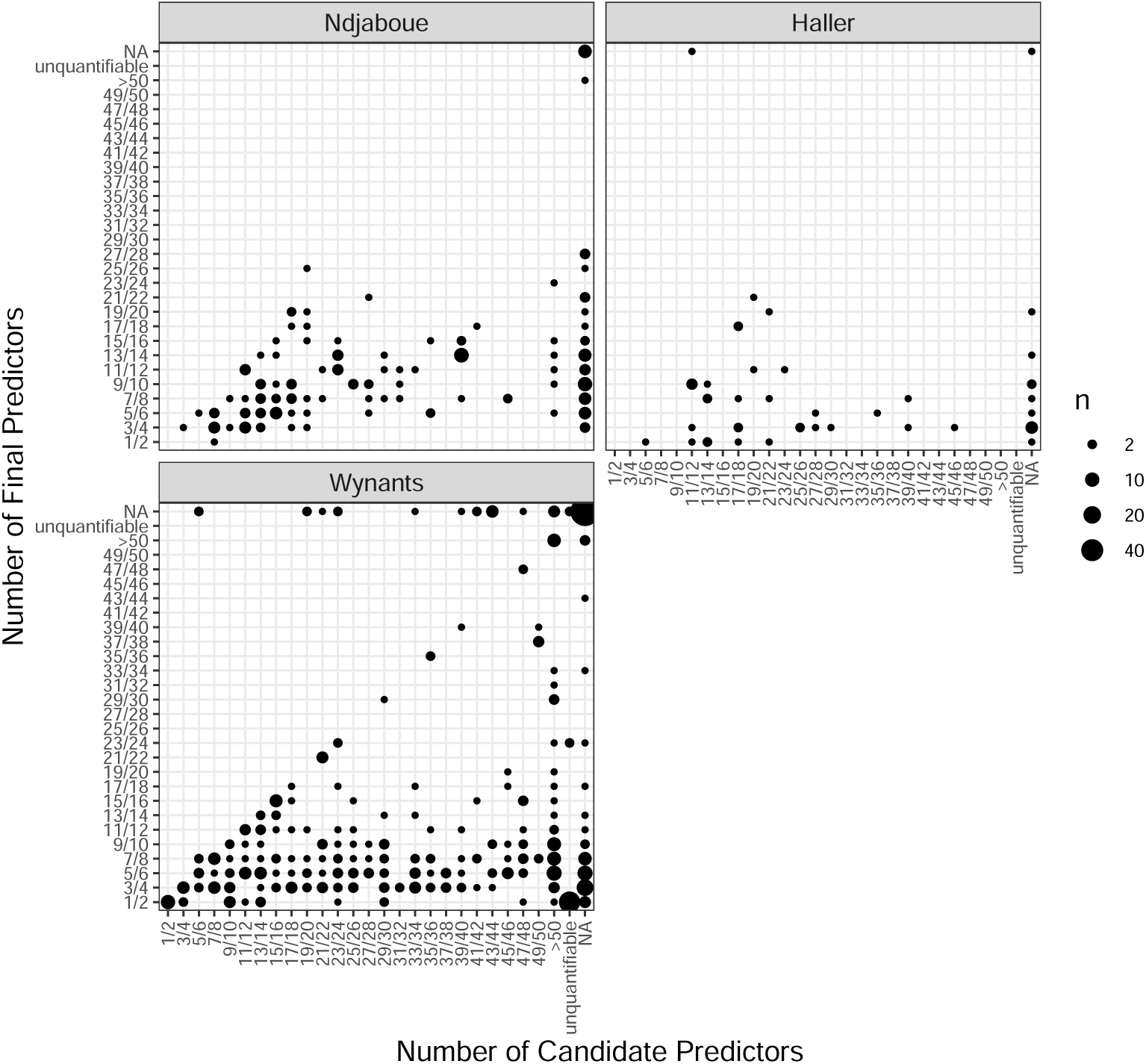
Number of candidate and final predictors, by review. The information is given here on a per-model basis. Note that the first row and the last column denotes ‘NA’, respectively. Note that in few cases the number of reported final predictors exceeded the number of reported candidate predictors (we speculate that this might be due to reporting errors, recoding of categorical variables, flexible modeling of numerical variables). The size of the individual dot (cf the value ‘n’ in the legend) corresponds to the number of overlapping points.

Information about the methods for predictor selection before or during modelling was not collected in all the reviews, and even for the reviews that collected this information, there were a lot of missing values. Regarding the pre-selection of predictors, out of the 586 models for which this information was in principle reported, selection based on univariate analyses was observed most frequently (192 models), followed by the use of all predictors (176 models) and a knowledge-based approach (32 models). The information about model selection during the model building was available even less often, with 159 models that used a stepwise approach, 84 times all variables were forced into the model (‘full-model approach’) and 41 models used a LASSO approach. Other methods were reported only for very few models.

#### 3.2.4 Number of models per paper

Seven reviews reported the number of models developed in each paper (77% of papers). Most papers presented the findings from one model (med=1, mean=1.8, IQR: 1 to 2, range: 1 to 28); one review included papers that reported considerably more models than the others (med=4, IQR: 2 to 5, Ogink [37]), the variability was larger in the most recent papers (Figure 3 and Supplementary file 1).

#### 3.2.5 Missing values

Five reviews collected information on missing values; in 54% of the papers no information about how missing data were handled was reported (Figure 5). Further, the quality of reporting did not increase over time. Complete case analysis was still the most common method, but the use of imputation methods became more frequent in recent years (mostly reported in Haller [41] and Sun [40], while complete case analyses were prevalent also in recent years in He [38] and Gade [42]). The review of Wynants [4] showed very similar results as observed in the papers from the other reviews analysed in this work and published in the same period (2020/21).

**Fig 5.**
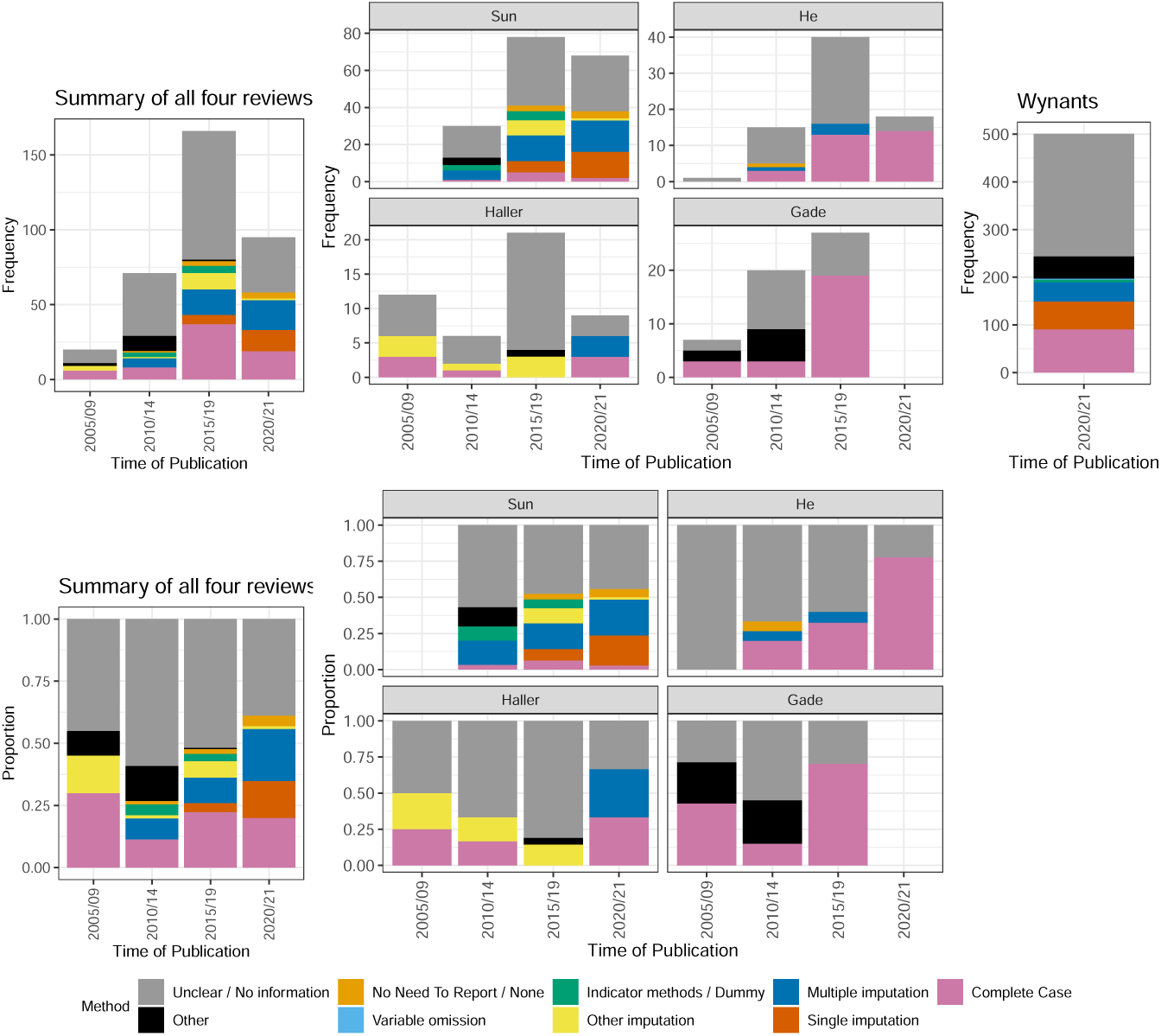
Methods used for handling missing data. The information was summarized on a per-model level using four reviews for which the information was available, overall (left) and stratified across reviews (right). Frequencies (top panels) and proportions (lower panels) are shown. Furthermore, we give frequencies for Wynants.

#### 3.2.6 Measures of predictive performance for internally validate models

One review (Ogink et al. [37]) did not collect information on model calibration, and four did not report classification measures (Haller [41], He [38], Ndjaboue [39], Sun [40]) (Table 3 and Figure 6), whereas discrimination was reported in all 8 reviews.

**Fig 6.**
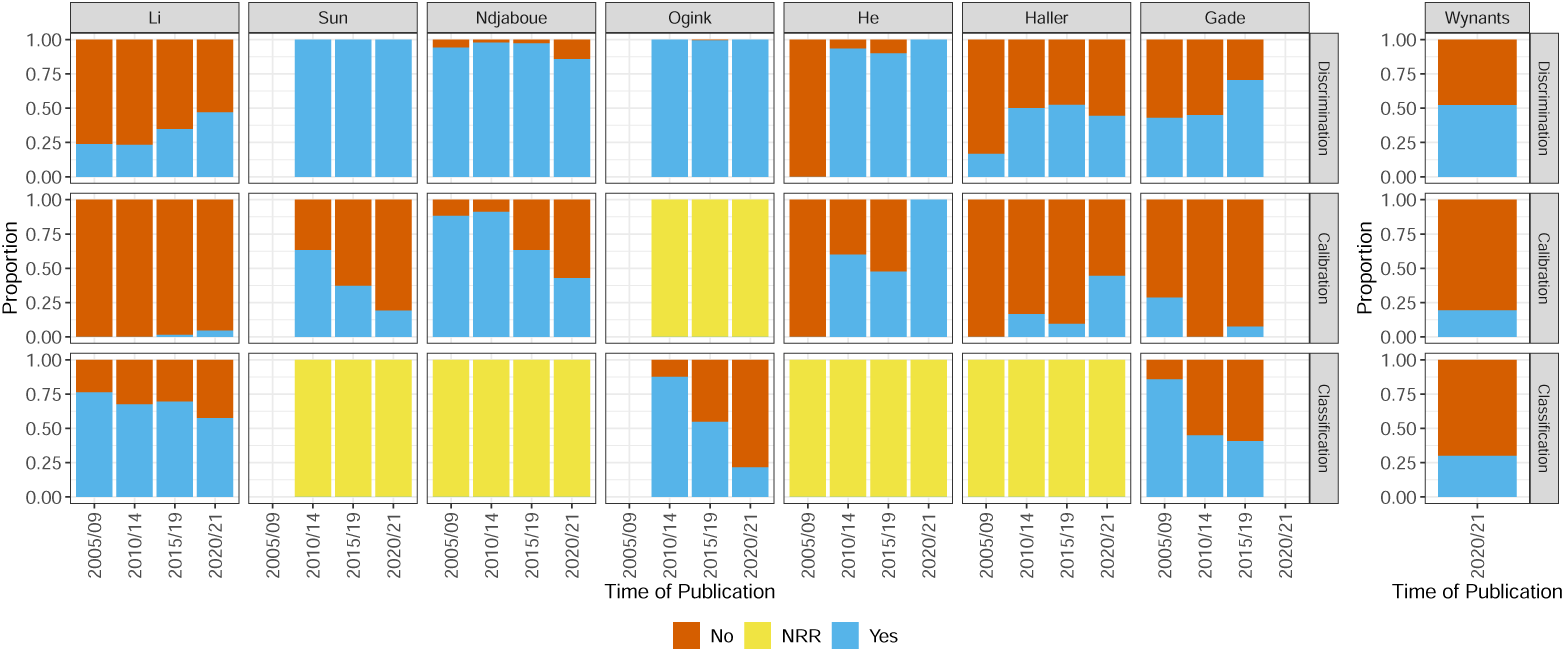
Summary of the reported measures. The display is stratified by review and time of publication and is given on a per-model level. The abbreviation ‘NRR’ stands for ‘not reported by the review’.

**Table 3.** Summary statistics by time of publication and by review. Number and percentage of papers where the model characteristic was reported. 2020/21*: Note that the papers summarized for the 2020/21 period do not include those from the Wynants review.

|  | Internal val. | External val. | Discrimination | Classification | Calibration |
| --- | --- | --- | --- | --- | --- |
| Overall | 575/887 (65%) | 170/831 (20%) | 563/887 (63%) | 285/647 (44%) | 221/831 (27%) |
| Time of publication |  |  |  |  |  |
| 2005/09 | 21/46 (46%) | 8/46 (17%) | 20/46 (43%) | 20/25 (80%) | 12/46 (26%) |
| 2010/14 | 82/148 (55%) | 27/141 (19%) | 98/148 (66%) | 47/72 (65%) | 47/141 (33%) |
| 2015/19 | 142/200 (71%) | 26/167 (16%) | 149/200 (74%) | 62/100 (62%) | 50/167 (30%) |
| 2020/21* | 82/125 (66%) | 14/109 (13%) | 87/125 (70%) | 42/82 (51%) | 30/109 (28%) |
| Review |  |  |  |  |  |
| Wynants | 248/368 (67%) | 95/368 (26%) | 209/368 (57%) | 114/368 (31%) | 82/368 (22%) |
| Li | 105/202 (52%) | 7/202 (3%) | 71/202 (35%) | 133/202 (66%) | 4/202 (2%) |
| Sun | 65/78 (83%) | 10/78 (13%) | 78/78 (100%) | - | 35/78 (45%) |
| Ndjaboue | 55/75 (73%) | 40/75 (53%) | 72/75 (96%) | - | 57/75 (76%) |
| Ogink | 47/56 (84%) | - | 55/56 (98%) | 25/56 (45%) | - |
| He | 36/52 (69%) | 12/52 (23%) | 47/52 (90%) | - | 34/52 (65%) |
| Haller | 12/35 (34%) | 5/35 (14%) | 18/35 (51%) | - | 7/35 (20%) |
| Gade | 7/21 (33%) | 1/21 (5%) | 13/21 (62%) | 13/21 (62%) | 2/21 (10%) |

Discrimination was reported for 63% of the papers, classification measures for 44%, calibration for 27%. Reporting of all three types of measures was rare (the co-occurrence of different measures is reported in the Supplementary file 1).

Reporting of discrimination improved over time (43% in 2005/09 period, 70% in 2020/21), but the reporting for classification and calibration did not (Table 3). There was considerable heterogeneity across reviews; for example, reporting of discrimination ranged from 35% (Li, [30]) to 100% (Sun, [40]), classification from 31% (Wynants [4]) to 62% (Gade [42]), and calibration from 2% (Li, [30]) to 76% (Ndjaboue, [39]) (Table 3 and Figure 6). In the Wynants review [4], the measures were reported less frequently compared to the papers from the other reviews analysed in this work and published in the same period (2020/21) (Table 3).

#### 3.2.7 Internal validation

All reviews collected information on internal validation of the included models. Overall, internal validation was performed in 575/887 (65%) of the papers. Findings varied across the reviews (Table 3), ranging from 33% in Gade [42] to 84% in Ogink [37]. Overall and in most reviews the reporting of internal validation increased with time (Figure 7). How internal validation was performed varied widely across reviews: the most commonly used methods for internal validation were cross-validation (very few other methods were observed in the reviews of Li and Ogink), bootstrap (which was common in He [38]) and split-sample methods, which are not recommended (or efficient) for regression based approaches [43], and were less common in the most recent periods and commonly used only in few reviews.

**Fig 7.**
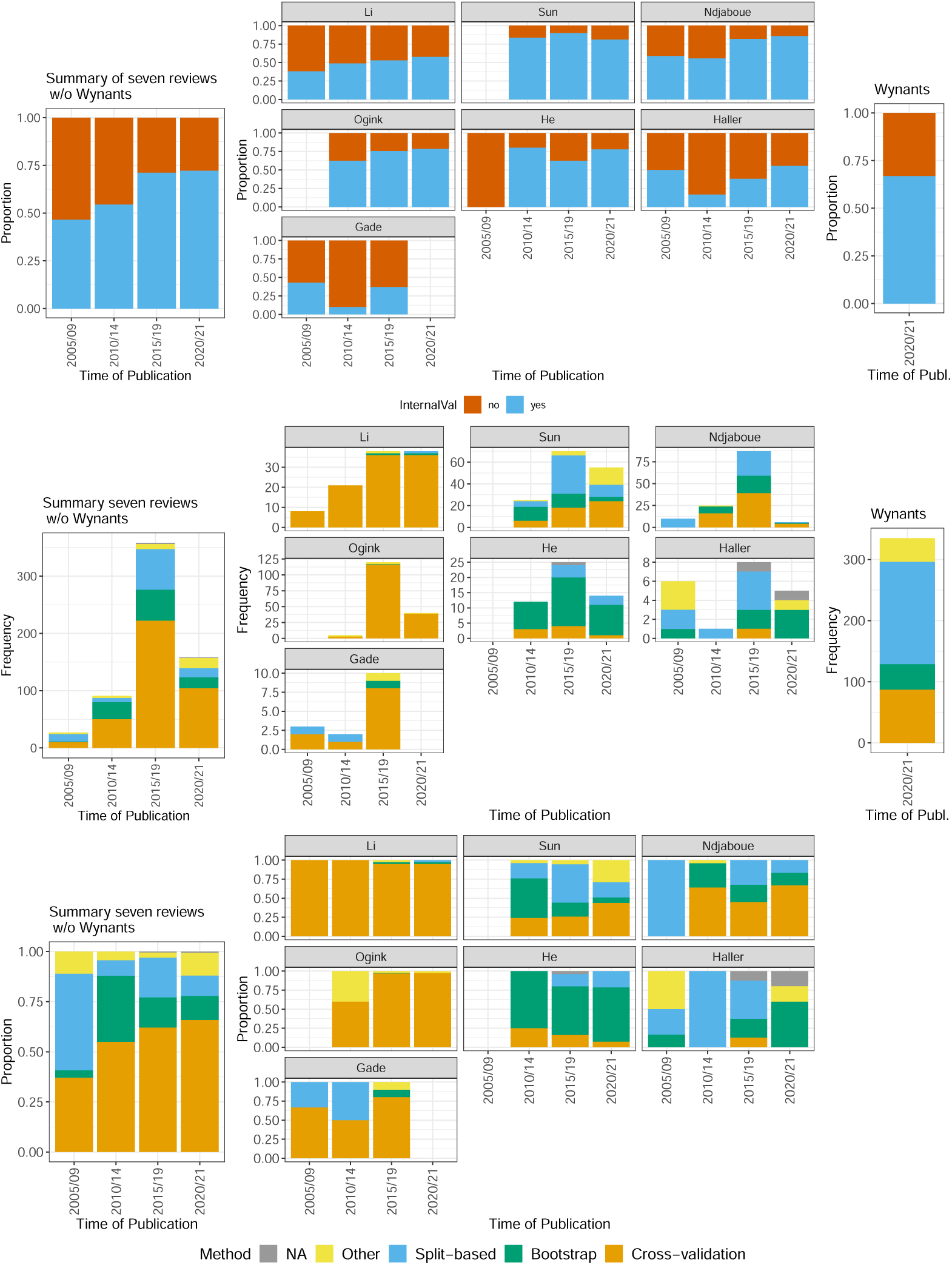
Top: Proportion of models that reported performing internal validation grouped by year of publication. Overall (left), by review (middle) and for Wynants (right). **Bottom: Methods used for internal validation, grouped by year of publication.** The analysis was restricted to the models for which using internal validation was reported. Overall (left), by review (middle) and for Wynants (right); absolute values (upper) and proportions (lower).

#### 3.2.8 External validation

One review did not collect information on external validation (Ogink [37]). Overall, external validation was reported for 170/887 (20%) of the papers. Reporting of external validation did not increase markedly with time (Figure 8, Table 3). The Wynants review had higher levels of external validation compared to the papers from the other reviews analysed in this work and published in the same period (2020/21) (26% vs 13% of papers).

**Fig 8.**
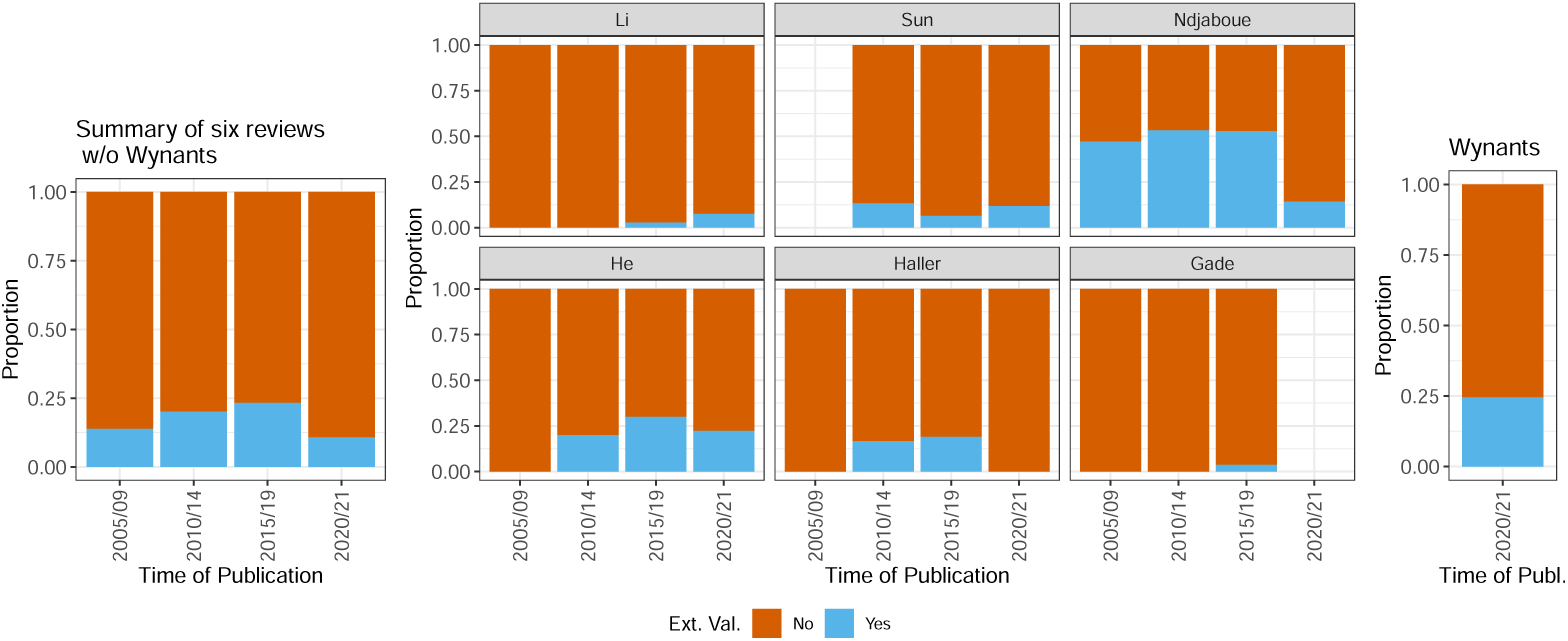
Proportion of models with reported external validation. The information is shown across all six reviews (left), stratified by review (middle), and for the Wynants review (right).

Reviews were heterogeneous also in terms of external validation, ranging from 5% of Gade [42] to 53% of Ndjaboue [39], in which the percentage was expected to be large due to the inclusion criteria used in that review, which included models with reported evidence of internal and/or external validation.

#### 3.2.9 Type of prediction model

All reviews collected information on the modelling approach to develop the prediction models. Overall, 51% of the papers used exclusively statistical approaches (e.g., regression based), 31% exclusively ML methods, 11% used both, and the information was unclear for 7% of the papers (Figure 9). Overall (excluding the Wynants [4] review), Cox regression was used in 21.6% of the models, (penalized) logistic regression in 16.5%, linear regression in 1.9%, neural networks in 17.9%, random forests in 5.9%, tree-based methods in 4.0%, SVM in 5.3%, and boosting in 4.5%. One review (He [38]) reported an almost exclusive use of the Cox model (Figure 10 from the paper), which was common also in Ndjaboue [39]. We did not retrieve the use of penalized Cox regression or of Cox regression with boosting.

**Fig 9.**
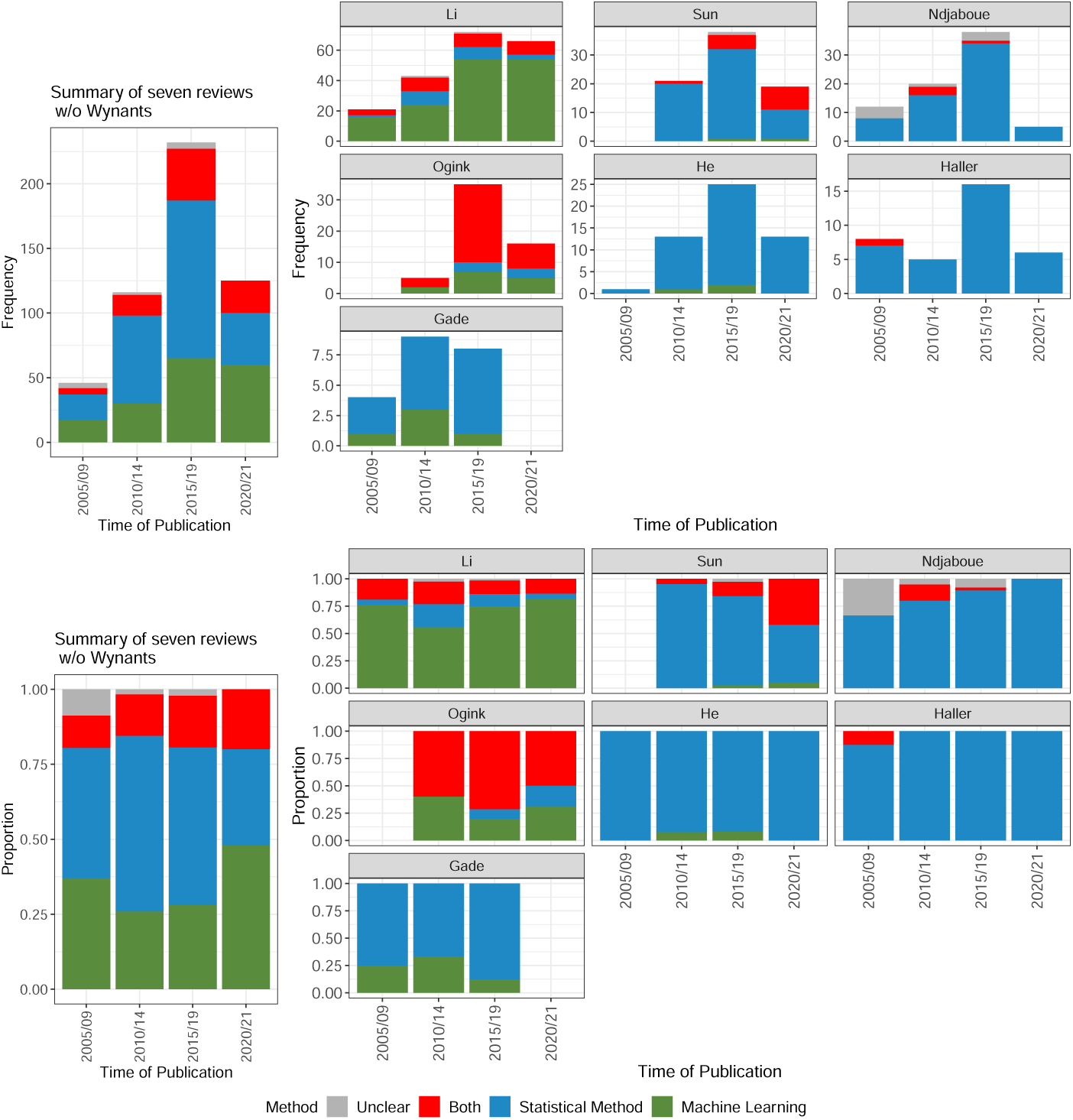
Categorized type of prediction model when counting purely per paper. This means, for each paper, the different models are considered and it is checked whether models corresponding to ‘Statistical Methods’ only, ‘Machine Learning’ methods only, or both are used.

**Fig 10.**
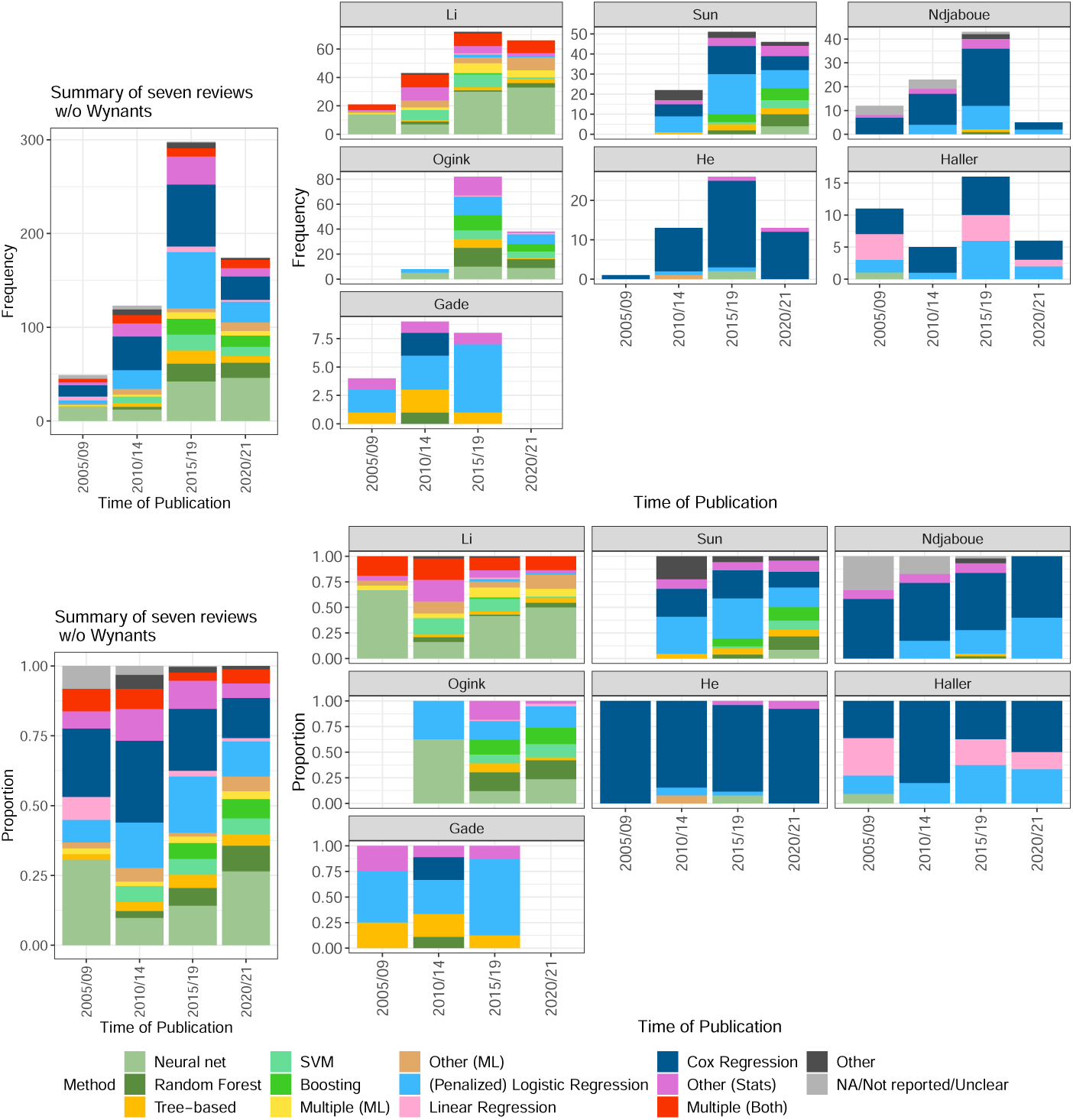
Different types of prediction models. The information is summarized across all reviews (left) and stratified by review (right). Frequencies are shown in the top figure, and proportions in the bottom. Counting was performed in a ‘per paper’ way, i.e. if, e.g., for one paper, three tree-based methods and 10 SVMs were reported, both would be counted only once.

**Fig 11.**
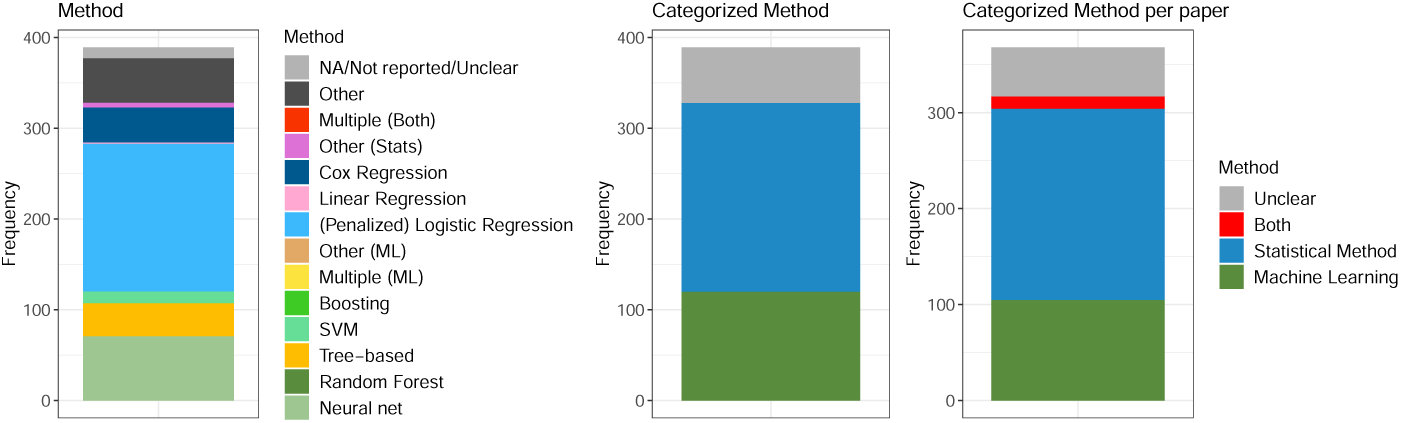
Types of prediction models for the Wynants review only. The plot on the left shows the individual methods, counted as explained for Figure 10. In the middle, these are summarized into the indicated categories. The plot on the right shows the result when considering each paper only once.

The results across reviews were inconsistent with no clear time trends (Figures 9 and 10, Table 4). Two reviews required the use of (at least one) ML method as an inclusion criterion for the selection of the papers (Ogink and Li, [30, 37]). In these two reviews the statistical methods were most rarely used exclusively (14 and 28%, respectively); nevertheless, the use of both ML and statistical methods was common in both reviews, especially in Ogink [37] (Figure 9). Two other reviews identified many papers that used exclusively ML methods (24% in Wynants [4] and 29% in Gade [42]), which was rare in the other four reviews.

**Table 4.**
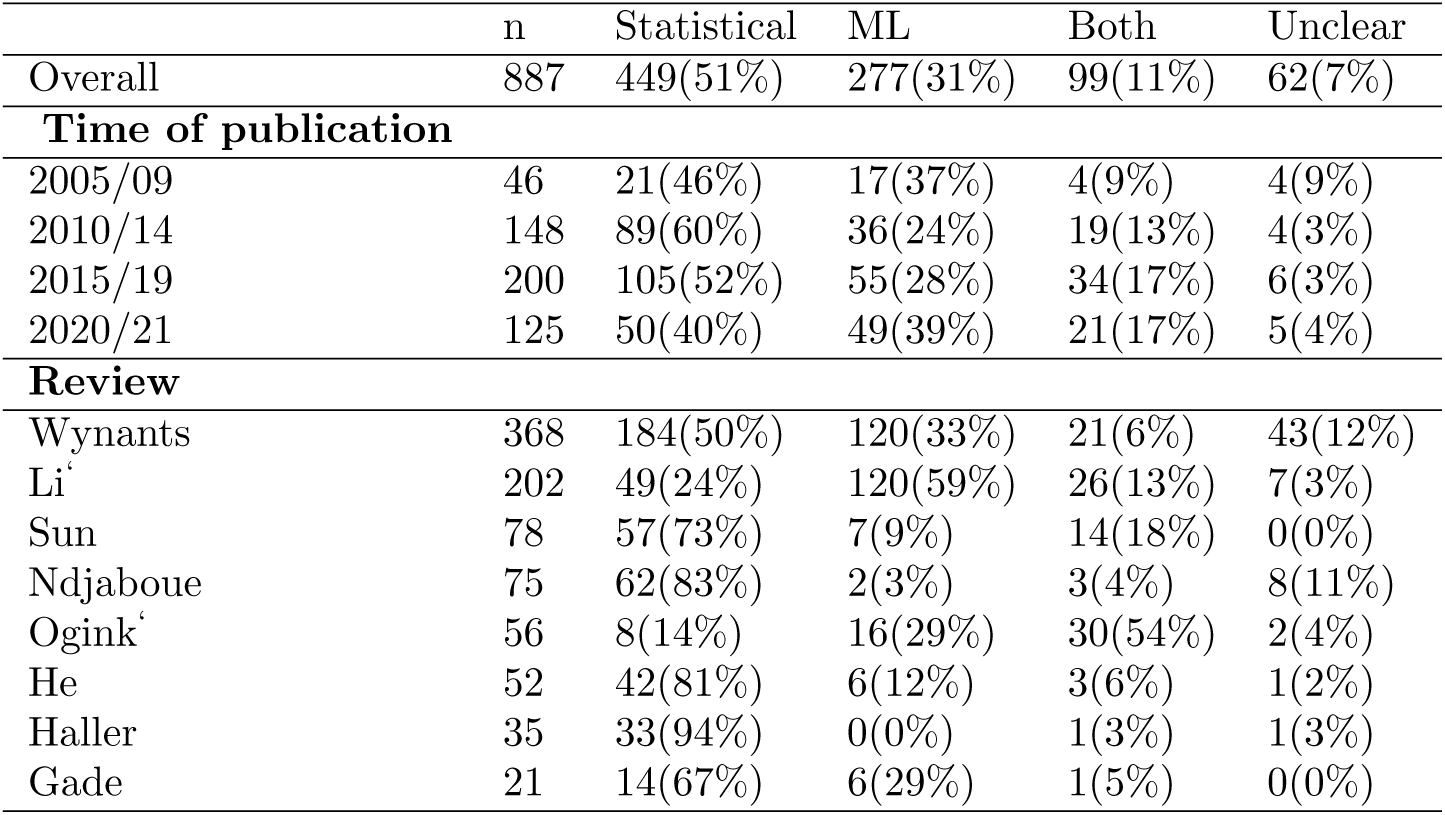
Number (percentage) of papers that used models that were statistical/ML/of both types/type was unclear, stratified by time of publication of the models and review. 2020/21*: note that the papers reported in the 2020/21 period do not include those from the Wynants review. ^*‘:reviewswithinclusioncriteriarelatedtotheuseof MLmethods*.^

One review indicated an increase in the use of both types of methods with time (Sun [40]), another an increase of the use of (penalized) logistic regression, but other clear time trends were not noticeable, both within individual reviews and overall. The large(r) percentage of ML methods observed in the 2020/21 period (39% vs 28% in the the 2015-20 period) seems attributable to the large influence of the data from the Li [30] review rather than to an overall increase, which is not observed within reviews. Moreover, the statistical methods were used more commonly in the Wynants review [4] than in the papers from the other reviews analysed in this work and published in the same period (2020/21) (50% vs 40%).

#### Comparisons by type of model

Papers that used exclusively statistical models used larger datasets and fewer candidate predictors compared to papers that used exclusively ML methods, which presented higher right-skewness in the distribution of the numerical variables (Table 5).

**Table 5.** Summary statistics by type of models. n (%) is the number (percentage) of papers for which the information was retrieved in the review.

| Model(s) | n (%) | Median | Mean | Range | IQR |
| --- | --- | --- | --- | --- | --- |
| **Number of study participants** |  |  |  |  |  |
| Statistical | 381 (85%) | 421 | 11,891 | 4 to 1,621,149 | 160 to 1475 |
| ML | 260 (94%) | 347 | 19,753 | 8 to 1,567,636 | 130 to 1071 |
| Both | 94 (95%) | 718 | 9246 | 20 to 246,405 | 192 to 5386 |
| Unclear | 49 (79%) | 360 | 65,176 | 20 to 3,041,551 | 128 to 1603 |
| **Number of outcome events** |  |  |  |  |  |
| Statistical | 292 (65 %) | 84 | 591 | 7 to 28,140 | 41 to 288 |
| ML | 166 (60 %) | 95 | 689 | 10 to 46,163 | 48 to 214 |
| Both | 49 (49 %) | 75 | 2572 | 5 to 74,661 | 44 to 268 |
| Unclear | 39 (63 %) | 98 | 1133 | 18 to 25,536 | 40 to 338 |
| **Number of candidate predictors** |  |  |  |  |  |
| Statistical | 225 (57 %) | 23 | 33 | 1 to 1224 | 14 to 37 |
| ML | 68 (28 %) | 33 | 289 | 7 to 15,000 | 24 to 49 |
| Both | 18 (18 %) | 22 | 23 | 2 to 45 | 12 to 32 |
| Unclear | 30 (48 %) | 13 | 33 | 2 to 166 | 9 to 43 |
| **Number of final predictors** |  |  |  |  |  |
| Statistical | 370 (82 %) | 6 | 12 | 1 to 488 | 4 to 10 |
| ML | 113 (41 %) | 6 | 19 | 2 to 618 | 3 to 13 |
| Both | 56 (57 %) | 9 | 88 | 2 to 3512 | 6 to 14 |
| Unclear | 39 (63 %) | 7 | 10 | 2 to 39 | 5 to 10 |
| **Number of models** |  |  |  |  |  |
| Statistical | 400 (89 %) | 1 | 2 | 1 to 10 | 1 to 2 |
| ML | 157 (57 %) | 1 | 1 | 1 to 8 | 1 to 1 |
| Both | 73 (74 %) | 4 | 4 | 1 to 28 | 2 to 5 |
| Unclear | 55 (89 %) | 1 | 1 | 1 to 4 | 1 to 2 |

The reporting of the measures varied widely by type of model, measures of discrimination and calibration were reported much more frequently when statistical models were used (Table 6), as were the number of candidate and final predictors (Table 4).

**Table 6.** Summary statistics of predictive performance measures by type of model(s).

|  | Discrimination | Calibration | Classification |
| --- | --- | --- | --- |
| Statistical | 334/449(74%) | 171/441(39%) | 96/255(38%) |
| ML | 124/277(45%) | 24/261(9%) | 129/262(49%) |
| Both | 80/99(81%) | 13/69(19%) | 41/78(53%) |
| Unclear | 25/62(40%) | 13/60(22%) | 19/52(37%) |

## 4 Discussion

The aim of this paper was to investigate any changes in prognostic model studies in the recent years. We used systematic reviews of prognostic models to evaluate if some important aspects in the development and reporting of models have changed considerably over time.

Our study was based on the findings of 8 systematic reviews, selected among those published in 2020-22 that reviewed more than 30 papers reporting development prognostic models, and provided sufficient publicly available information for the re-analysis of most of the information guided by the CHARMS checklist. We re-analyzed the findings from 887 papers and 1448 models.

The findings from our study, based on these 8 reviews, show that the changes in prediction modeling are not as substantial as it might have been anticipated. Some of the key findings of our paper are: models did not become substantially bigger over time (with respect to the number of variables); within each review we did not observe an increase of the use of ML methods over time; discrimination assessments are still much more popular than calibration assessments; there is an indication of a trend towards increasingly following guidelines (e.g. with respect to performing/reporting internal validation, and using resampling methods instead of sample splitting).

We observed that the number of study participants (and outcome events) increased in time, the substantial increase in the 2015/19 period was followed in 2020/21 by a further increase, due to the presence of extremely large studies (e.g., using registry studies), but the central tendency (median) remained unchanged, indicating that only few studies contribute to the average changes. A similar pattern was observed for the mean number of final predictors, which substantially increased in the 2015/19 period, but for which the median values remained stable over time. These findings are somewhat surprising given that the amount of available data (and thus, the number of available predictors) has strongly increased during the past decade. It is debatable as to why this is the case; possible reasons could be that some unreported pre-selection of candidate predictors is being performed, thus diminishing visible increase in the number of predictors, that simpler models that use fewer predictors enable greater usability, interpretability, and transferability, and are thus still preferred, or that there is a time lag that prevents the detection of such increase, yet. Another possible explanation is that our review undersampled prognostic models based on imaging, as only two reviews included some prediction models based on imaging. An interesting finding, based only on the data from the Wynants review [4], was that imaging models had, as expected, more candidate predictors, but ended up using fewer predictors than the other models. This might indicate that the higher complexity of these data might play a crucial role especially in model development, with the dangers related to overfitting, and in the crucial need for proper internal and external validation.

Some recommendations contained in methodological guidance are seemingly increasingly being followed more closely. For example, the use of internal validation increased with time (e.g., bootstrapping), whilst relying on split-sample apprpaches became less commonly used. The use of external validation remained rather stable in time; however, our paper investigated only external validation contextually to model development, and therefore underestimates the proportion of models that are eventually externally validated (in subsequent papers). With time, the reporting of discrimination measures improved, while it did not for calibration and classification measures. We decided to report classification measures, as they were reported in 4 reviews and for more than 40% of the papers. However, their usefulness in assessing the performance of predictive models is not generally accepted [44] and we do not advocate that their reporting should be more common. In our data the use of ML learning methods was common and somewhat increased with time; however, generally this was not observed within reviews, where the type of model used remained rather stable in time. The prognostic modeling does not seem to be overwhelmed by ML models, nor by being based on extremely large data sets. Most of the research is still conducted using moderately sized data sets, both in terms of number of study participants and of number of variables.

The comparison between ML and statistical models indicated that the median number of study participants was smaller for ML models, similarly as observed in [45], but they had a larger arithmetic mean, and a somehow larger number of outcome events. The number of candidate predictors was larger (both in terms of median and mean), while the median number of used variables was very similar. Statistical and ML studies differed substantially in terms of reporting of model performance measures, especially calibration was very poorly reported for ML models.

Overall, we observed a considerable heterogeneity in the results from different reviews, indicating that the different medical fields might present very different characteristics in the development and reporting of prognostic models, and in the data being used. For example, the results from the comprehensive review on COVID-19 [4] differed in several aspects from those based on papers from the same period included in the other reviews. Time pressure to derive models intended to help handling the COVID-19 crisis is one potential explanation, but it may also indicate the need to consider different fields of application for a better understanding of the overall trends and characteristics of prognostic modeling. Moreover, a large group of experienced reviewers were involved in the COVID-19 project.

Many of the findings from the review surveying the prediction papers from 2008 [5] are still relevant today: reporting of practices are not consistently followed, external validation is still very uncommon, as is the evaluation of calibration. Some measures are still not reported in the majority of papers, and some reviews do not collect all the relevant information. The TRIPOD statement [22] was mentioned in 6 out of the 8 considered reviews, and three reviews considered the aspects of the TRIPOD statement in detail with respect to the papers they reviewed.

Our study had several limitations. The papers included in our study are just a part of the many that are being developed and published, which are not currently included in systematic reviews. The findings of some big reviews were not available as raw data, and some important information was missing (by design) also from the reviews that we included. For example, one review did not report the number of study participants and outcome events, four did not report the number of candidate predictors; consequently, we could not fully explore the number of outcome events per variable, to gauge the risk of overfitting of the included prediction models. For the same reason we did not attempt to compare high and low-dimensional prediction models.

The implementation of systematic reviews should be consistent with the guidelines that are available to increase the usefulness of their findings [27], which would be further improved if the raw data were made routinely publicly available.

A further limitation consisted in some characteristics of the reviews that we considered. For example, one review included only papers that reported some type of internal or external validation, inflating the number of such papers in our analyses. Two of the selected reviews included only papers that reported the use of at least one machine learning method, which could inflate our estimate of the frequency of the use of ML methods. Nevertheless, our further manual categorization of the methods being used indicated that these reviews included many models that were developed using statistical methods. Similarly, a review of machine learning based clinical prediction models published in 2019 in the field of oncology, found that regression-based models (such as logistic or Cox regression) were categorized as ML methods by the authors very often, and constituted about a third of the prognostic models that they reviewed [45]. The study design of the papers was not reported in most of the reviews; when reported, observational studies where the vast majority; therefore, studies based on registries and on randomized clinical trials might be underrepresented. Also prognostic modeling based on imaging data might be underrepresented in our review. To the best of our knowledge, only two of the selected reviews included at least partly prognostic models based on imaging data, and a direct comparison between models using images and other models was not feasible.

## Data Availability

Data will be available as Supplementary table (included in the submission)

## Supporting information

**Supplementary file 1 Supplementary file** The file includes additional description of the methods and results not presented in the main paper.

**Supplementary table 1 Data table** The file includes the raw data used in this paper, where the information about papers/models reviewed in each review were retrieved and harmonized for the analyses presented in this paper.

## Acknowledgments

This work was developed as part of the international initiative of Strengthening Analytical Thinking for Observational Studies (STRATOS). The objective of STRATOS is to provide accessible and accurate guidance in the design and analysis of observational studies (http://stratos-initiative.org/).

The members of the Topic Group 9: High-dimensional data of the STRATOS initiative are: Federico Ambrogi, Axel Benner, Harald Binder, Anne-Laure Boulesteix, Riccardo De Bin, Kevin Dobbin, Roman Hornung, Lara Lusa, Lisa M. McShane, Stefan Michiels, Eugenia Migliavacca, Joerg Rahnenfuehrer, Willi Sauerbrei and Martin Treppner.

L.L. was partially supported by ARRS research program P3-0154. W.S. was partially supported by grant SA580/10-1 from the German Research Foundation (DFG).

## Notes

### Competing Interest Statement

The authors have declared no competing interest.

### Funding Statement

Yes

### Author Declarations

N/A Approval not necessary, as it is a re-analysis of systematic review papers.

